# Plasma (1→3) β-d-glucan levels are associated with host inflammatory responses and predict adverse clinical outcomes in critical illness

**DOI:** 10.1101/2020.06.11.20128264

**Authors:** Georgios D. Kitsios, Daniel Kotok, Haopu Yang, Malcolm Finkelman, Yonglong Zhang, Noel Britton, Rui Guo, John W. Evankovich, William Bain, Faraaz Shah, Yingze Zhang, Panayiotis V. Benos, Bryan J. McVerry, Alison Morris

## Abstract

**Background:** The fungal cell-wall constituent (1,3)-β-d-glucan (BDG) is a pathogen-associated molecular pattern (PAMP) that can stimulate innate immunity. We hypothesized that BDG from colonizing fungi in critically-ill patients may translocate into the systemic circulation and thus be associated with host inflammatory responses and outcomes.

**Methods:** We enrolled 453 mechanically-ventilated patients with acute respiratory failure with no evidence of invasive fungal infection (IFI). From serial plasma samples, we measured BDG, innate immunity and epithelial permeability biomarkers. From lower respiratory tract and stool samples we quantified bacterial and fungal DNA load using culture-independent techniques.

**Results:** A positive BDG test (>60pg/ml) at baseline was detected in 19% of patients. BDG levels were significantly associated with markers of innate immunity (interleukin-6, tumor necrosis factor receptor-1 and procalcitonin), epithelial barrier disruption (receptor for advanced glycation end-products and fatty-acid binding protein-2, for lung and gut respectively) and with higher probability of classification in an adverse prognosis hyperinflammatory subphenotype (all p<0.05). No differences in fungal or bacterial DNA load were found by BDG test positivity. Positive BDG testing was associated with higher incidence of acute kidney injury, fewer ventilator free days and worse 30-day survival (adjusted p<0.05). Patients with positive BDG test on follow-up sampling (>3 days from intubation) had higher mortality than patients with persistently negative test on follow-up (p<0.05).

**Conclusions:** This is the first study to demonstrate the prognostic role of BDG in critically ill patients with no evidence of IFI. Translocation of BDG into systemic circulation may contribute to inflammation and clinical outcomes.

**Funding support:** National Institutes of Health [K23 HL139987 (GDK); U01 HL098962 (AM); P01 HL114453 (BJM); R01 HL097376 (BJM); K24 HL123342 (AM); U01 HL137159 (DVM, PVB); R01 LM012087 (DVM, PVB); R01 HL142084 (JSL); R01 HL136143 (JSL); F32 HL137258 (JWE); F32 HL142172 (WB); K08 HS025455 (IJB); K23 GM122069 (FS)].

## Introduction

Dysregulation of innate immunity is a central pathogenetic feature of the heterogeneous syndromes of sepsis and the acute respiratory distress syndrome (ARDS) (1, 2). In several independent patient populations, patient subgroups with elevated plasma levels of inflammatory cytokines and biomarkers of tissue injury (classified as a *hyperinflammatory subphenotype*) exhibit worse organ dysfunction and clinical outcomes compared to their *hypoinflammatory* counterparts (3–5). Such biologic subphenotyping of critically-ill patients offers improved risk prediction over stratification with clinical variables alone, and presents new opportunities for targeted pharmacologic interventions (6). However, the molecular mechanisms of aberrant innate immunity stimulation and resultant tissue injury remain largely undefined.

Inter-patient variability of endogenous microbiota may explain the observed differences in host-responses. Many of the predictive biomarkers for the detrimental hyperinflammatory subphenotype (e.g. interleukins [IL]-6 and −8, soluble tumor necrosis factor receptor-1 [TNFR1], procalcitonin and receptor for advanced glycation end-products [RAGE]) (3, 4) reflect canonical pathways of immune cell stimulation from pathogen and damage associated molecular patterns (PAMPs and DAMPs, respectively) (7, 8). PAMPs include microbial nucleic acids as well as cell wall components, such as lipopolysaccharide of gram-negative bacteria and 1-3-beta-D-glucan (BDG) from fungi (8, 9). Recent culture-independent studies with bacterial DNA sequencing have uncovered important roles of intestinal and respiratory bacterial communities in the evolution and outcome of critical illness (10–13). However, very few studies have performed in depth examinations of the fungal communities (mycobiome) in critical illness (14) and little is known about how fungal PAMPs may interact with the innate immunity system in the critically-ill host.

Epidemiologic evidence suggests that host-fungal interactions at different mucosal surfaces are clinically important (15). In mechanically-ventilated immunocompromised patients, airway colonization by *Candida* species portends worse outcome (16), and in patients with suspected ventilator-associated pneumonia, *Candida* species in respiratory tract secretions have been associated with increased mortality and levels of IL-6 and procalcitonin (17, 18). *C*.*albicans* overgrowth in the human gut occurs in the setting of antibiotic-treated sepsis, raising the risk for secondary candidemia (19, 20). The respiratory and intestinal tracts in critical illness contain high levels of the fungal PAMP BDG. BDG is recognized by mucosal innate immune cells through the dectin-1 receptor and stimulates local inflammatory responses (21). It can also exert systemic effects on host inflammation via translocation into the bloodstream, in the setting of disruption of mucosal epithelial integrity from various insults, such as hypoxemia, ischemia or direct pathogen invasion (22). Preliminary observations have suggested that higher levels of circulating BDG are associated with worse clinical outcomes in patients in the intensive care unit (ICU) (23), but evidence has been limited. BDG levels are routinely checked in the diagnostic work-up of critically-ill patients with clinical suspicion of invasive fungal infection (IFI) (24), but whether BDG could be used as a clinical biomarker of fungal PAMP translocation and its contribution to the development and perpetuation of hyperinflammatory host-responses in critical illness is unknown.

In this study, we demonstrate that approximately 20% of critically-ill patients without IFI have elevated plasma BDG levels (as defined by a positive BDG test). Patients with a positive plasma BDG test had higher levels of circulating biomarkers of innate immunity and tissue injury, were more likely to be classified as the adversely prognostic hyperinflammatory subphenotype, and had worse clinical outcomes independent of other prognostic factors. Exploratory analyses for the potential source of BDG e.g. the intestinal or respiratory tract, suggested the potential presence of heterogeneous mechanisms underlying fungal PAMP translocation in critical illness.

## Results

### Study population

We analyzed data from 453 mechanically-ventilated patients with acute respiratory failure, who were consecutively enrolled from ICUs at the University of Pittsburgh Medical Center (UPMC) (3, 25). We excluded 11 patients with a clinical diagnosis of IFI (24) because we wanted to examine BDG as a fungal PAMP in a broad ICU population and not in the context of IFI as a diagnostic test. However, inclusion of patients with IFI in sensitivity analyses did not impact the observed associations and outcomes. Characteristics of included patients are shown in Table 1.

**Table 1:** Baseline characteristics of enrolled subjects, stratified by positive and negative BDG testing. P-values from nonparametric tests comparing patients with a negative (≤60 pg/ml) and positive (>60 pg/ml) BDG result are shown in bold when significant (p<0.05).

|  | Overall | Negative | Positive | p |
| --- | --- | --- | --- | --- |
| n | 453 | 368 | 85 |  |
| Age (median [IQR]) | 58.3 [46.0, 67.1] | 57.4 [46.5, 67.5] | 59.6 [43.7, 66.2] | 0.90 |
| Male (%) | 244 (53.9) | 196 (53.3) | 48 (56.5) | 0.68 |
| BMI (median [IQR]) | 28.4 [24.8, 35.7] | 29.1 [24.8, 36.5] | 27.5 [25.2, 32.2] | 0.08 |
| SOFA Score (median [IQR]) | 7 [4, 9] | 6 [4, 9] | 8 [5, 10] | **0.01** |
| P:F Ratio (median [IQR]) | 164 [117, 205] | 165 [117, 206] | 164 [120, 205] | 0.81 |
| History of COPD (%) | 108 (23.8) | 90 (24.5) | 18 (21.2) | 0.62 |
| History of diabetes mellitus (%) | 150 (33.1) | 121 (32.9) | 29 (34.1) | 0.93 |
| History of immunosuppression (%) | 88 (19.4) | 66 (17.9) | 22 (25.9) | 0.13 |
| Group classification (%) |  |  |  | 0.96 |
| Acute on chronic respiratory failure | 24 (5.3) | 19 (5.2) | 5 (5.9) |  |
| ARDS | 120 (26.5) | 95 (25.8) | 25 (29.4) |  |
| At Risk for ARDS | 172 (38.0) | 142 (38.6) | 30 (35.3) |  |
| Control - Airway Protection | 67 (14.8) | 54 (14.7) | 13 (15.3) |  |
| Control - CHF | 34 (7.5) | 29 (7.9) | 5 (5.9) |  |
| Other | 36 (7.9) | 29 (7.9) | 7 (8.2) |  |
| BDG (median [IQR]) | 26 [15, 49] | 21 [13, 33] | 95 [76, 227] | **<0.001** |
| Direct injury, "pulmonary" ARDS (%) | 209 (46.1) | 168 (45.7) | 41 (48.2) | 0.76 |
| Beta-lactam on study day 1 (%) | 340 (75.7) | 270 (74.2) | 70 (82.4) | 0.15 |
| Topical antifungal on study day 1 (%) | 13 (2.9) | 9 (2.5) | 4 (4.7) | 0.45 |
| Systemic antifungal on study day 1 (%) | 4 (0.9) | 2 (0.5) | 2 (2.4) | 0.34 |
| Positive respiratory culture (%) | 105 (23.2) | 84 (22.8) | 21 (24.7) | 0.82 |
| Incident shock (%) | 226 (49.9) | 178 (48.4) | 48 (56.5) | 0.22 |
| Bacteremia (%) | 44 (9.7) | 37 (10.1) | 7 (8.2) | 0.76 |
| Hyperinflammatory subphenotype (%) | 127 (28.0) | 89 (24.2) | 38 (44.7) | **<0.001** |

P-values from nonparametric tests comparing patients with a negative ( $\leq 60$ pg/ml) and positive ( $> 60$ pg/ml) BDG result are shown in bold when significant ( $p < 0.05$ ). BMI: Body mass index, SOFA Score: Sequential organ failure assessment score, P:F Ratio: Ratio of partial pressure of arterial oxygen and fraction of inspired oxygen, COPD: Chronic obstructive pulmonary

### Distribution of plasma BDG levels in critically-ill patients

At baseline, 85 patients (19%) had positive BDG defined as greater or equal to the conventional threshold of 60 pg/ml (Figure 1) (24). Patients with positive BDG levels (median 95, interquartile range [IQR] 76-227 pg/ml) had similar distribution of baseline clinical characteristics as patients with negative BDG (median 26, IQR 15-49 pg/ml), with the exception of higher sequential organ failure assessment (SOFA) scores in the positive group (p=0.01, Table 1). Similar results were obtained when we used a more stringent threshold of BDG positivity (80 pg/ml, Table S1). Although beta-lactam antibiotics have been previously implicated in false-positive BDG detection (26, 27), we did not find any differences in BDG levels between patients receiving beta-lactam antibiotics, or not, at the time of sampling. A small proportion of patients (2.9%) receiving topical antifungals for either oral thrush or skin candidiasis (clotrimazole troche or nystatin) had significantly higher BDG levels compared to those not receiving these medications (n = 13 [2.9%], p=0.03).

**Figure 1:**
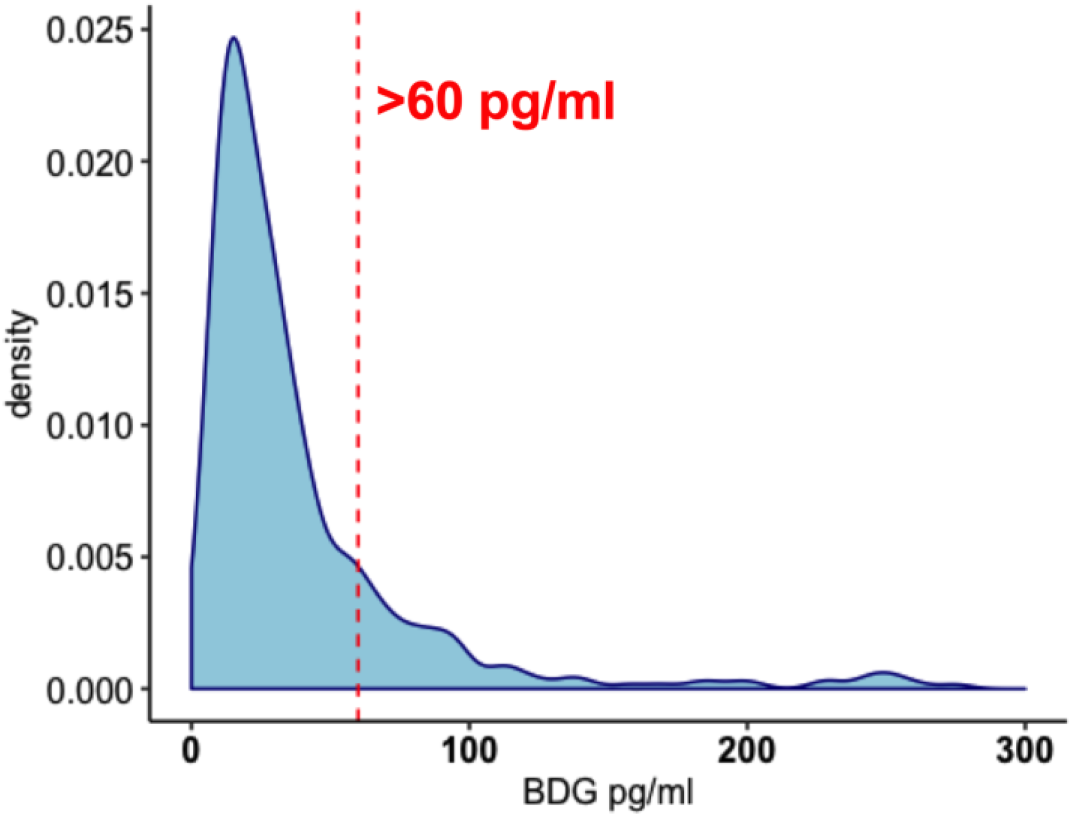
Density histogram of measured 1,3-beta-D-glucan (BDG) plasma levels in a cohort of 453 critically ill patients. 19% of patients had BDG levels >60pg/ml which is the conventional threshold for a positive BDG test in the diagnostic workup of invasive fungal infection.

### Plasma BDG correlates with host-response biomarkers and the hyperinflammatory subphenotype

We systematically profiled host-responses by measuring plasma biomarkers in pathways of innate immunity (IL-6, IL-8, IL-10, TNFR1, fractalkine, suppression of tumorigenicity-2 [ST-2]), alveolar epithelial injury (RAGE), endothelial injury (angiopoeitin-2) and response to bacterial infection (procalcitonin and pentraxin-3) (3). BDG levels at baseline positively correlated with levels of all measured biomarkers (p<0.05), with seven of these correlations remaining significant after adjustments for multiple testing (Figure 2A). We synthesized these host-response profiles into a parsimonious regression model for classification into a hyperinflammatory (28%) vs. a hypoinflammatory subphenotype (72%) based on latent class analyses as previously described (3). Hyperinflammatory patients had significantly higher mean BDG levels than hypoinflammatory ones (39 [23-68] vs. 23 [14-39] pg/ml, p<10^−6^, Figure 2B).

**Figure 2:**
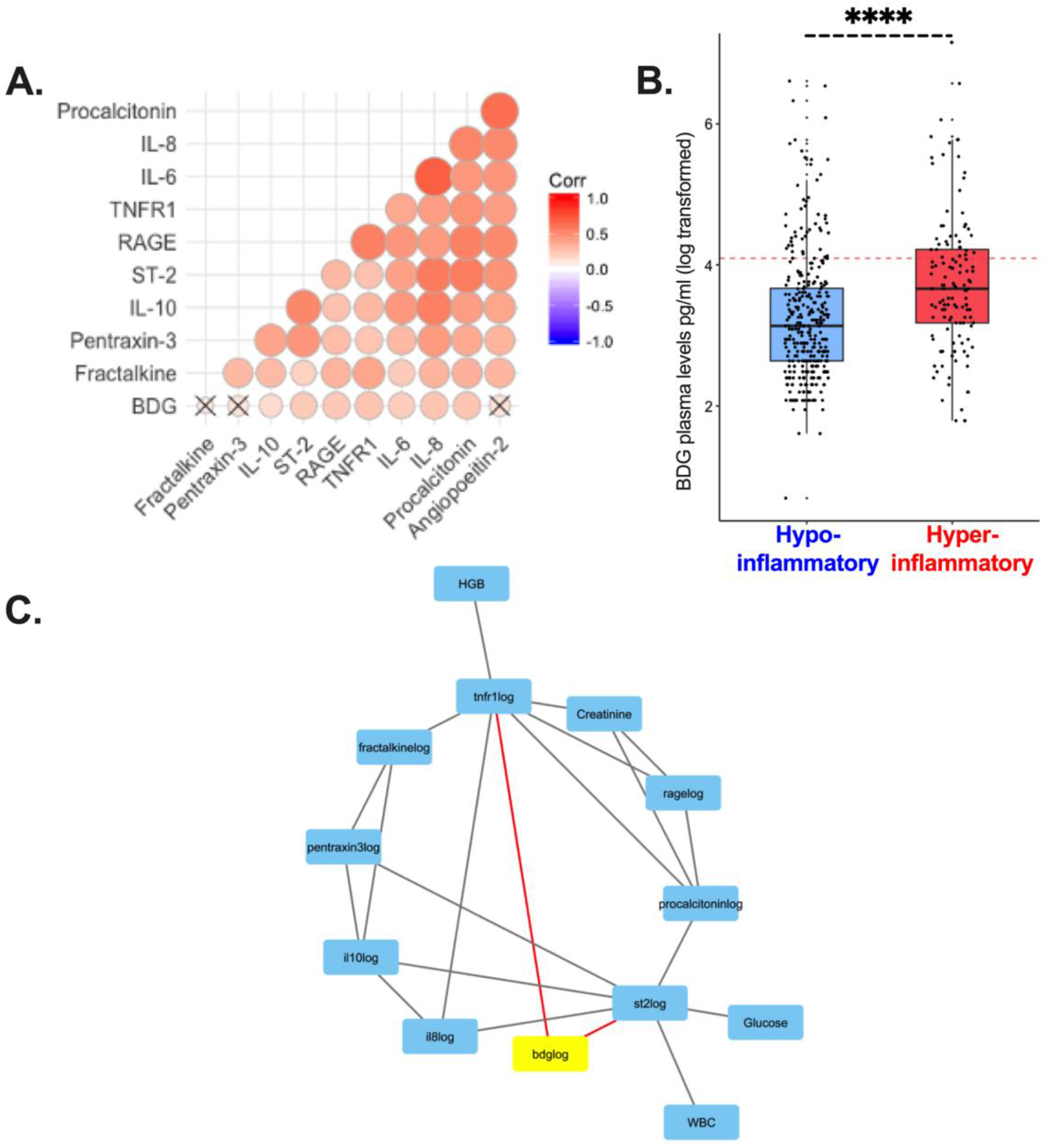
BDG levels are associated with inflammatory host-responses. A. Correlogram of 10 host-response biomarkers and BDG levels. Effect sizes of positive correlations are shown with variations of red, and statistical significance by size of circles. Comparisons were adjusted for multiple testing with the Benjamini-Hochberg method. Non-significant correlations are marked with X. B. Patients assigned to the hyperinflammatory subphenotype had significantly higher levels of BDG (p<0.0001). C. Probabilistic Graphical Model analysis demonstrating first and second neighbors of the BDG variable, when considered in conjunction with 32 clinical and biomarker variables. Two first neighbors were identified: TNFR1 and ST-2. Abbreviations: IL-6: Interleukin-6; IL-10: Interleukin-10; TNFR-1: Tumor necrosis factor receptor-1; Ang-2: Angiopoietin-2; ST-2: Suppression of tumorigenicity-2; RAGE: Receptor for advanced glycation end products.

### Network analyses reveal association of BDG with key elements of the hyperinflammatory response

To comprehensively examine for causal associations between baseline clinical variables, biomarkers and BDG, we utilized the Probabilistic Graphical Models (PGM) framework (3, 28, 29). From a total of 32 variables, only two were directly linked with BDG (i.e. first neighbors in PGM): TNFR1, a predictive biomarker for the hyperinflammatory subphenotype, and ST-2, the soluble receptor of IL-33 that functions as an alarmin released by respiratory epithelia during acute lung injury (30). Both TNFR1 and ST-2 were further linked to additional components of the hyperinflammatory response (Figure 2C).

### Patients with higher BDG levels had worse clinical outcomes

BDG levels analyzed as a continuous variable were associated in regression models with higher incidence of acute kidney injury (AKI), fewer ventilator-free days (VFDs) and higher 30-day mortality (all p<0.05, Table 2). These associations with clinical endpoints remained significant after adjustments for other important predictors of outcome in critical illness (age, SOFA scores and hyperinflammatory subphenotype) as well as potential confounders of BDG measurement (beta-lactam antibiotics that have been associated with false positive BDG and batch of BDG measurement). Patients with positive BDG testing had worse 30-day survival (adjusted hazard ratio: 1.37, 95% confidence interval 1.13-1.67, p<0.001) compared to patients with negative BDG (Figure 3).

**Table 2:** Associations between BDG levels and clinical outcomes. Outcomes of BDG-positive and negative patients are shown in the first two columns. P-values for the statistical significance of the association between BDG levels and each outcome (Logistic regression models for acute kidney injury and 30-day mortality; zero-inflated negative binomial regression for ventilator-free days) are shown for unadjusted and adjusted analyses (adjustments for age, sequential organ failure assessment score, hyperinflammatory subphenotype assignment, beta-lactam antibiotics and batch of BDG measurement). Statistically-significant associations are highlighted in bold.

|  | BDG-positive | BDG-negative | Unadjusted p-value | Adjusted p-value |
| --- | --- | --- | --- | --- |
| Acute Kidney Injury, n (%) | 63 (74%) | 237 (64%) | **0.008** | 0.10 |
| Ventilator-free days, median (IQR) | 16 (0-23) | 19 (0-24) | **<0.0001** | **0.005** |
| 30-day mortality, n (%) | 34 (40%) | 86 (23%) | **0.001** | **0.01** |

**Figure 3:**
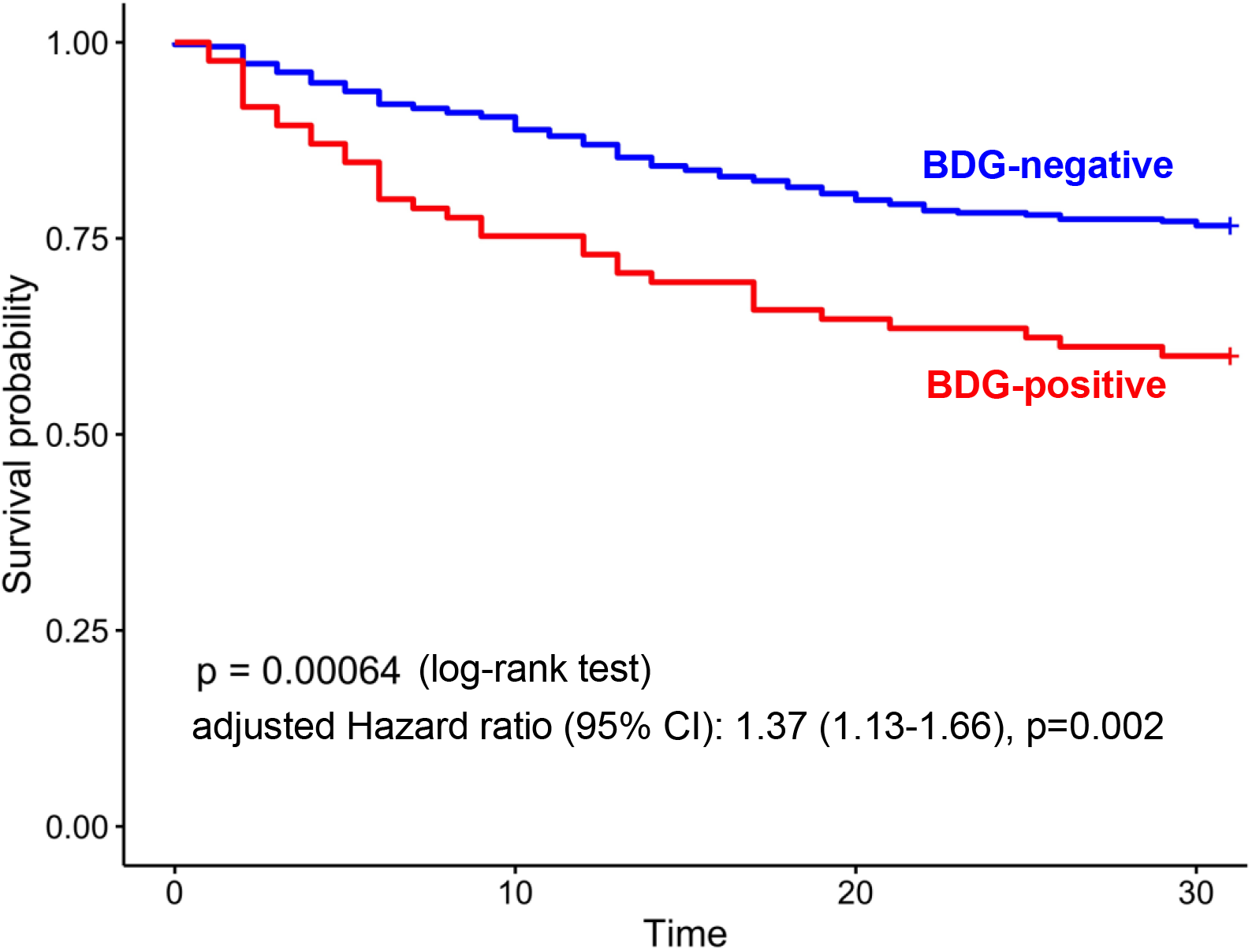
Patients with a positive BDG test had worse 30-day survival compared to patients with negative BDG test. Kaplan-Meier curve for 30-day survival. Hazard ratio adjusted for age, sequential organ failure assessment score, hyperinflammatory subphenotype assignment, beta-lactam antibiotics and batch of BDG measurement.

### BDG is associated with markers of epithelial barrier disruption in the respiratory and intestinal tract

Given the clinically significant prognostic associations of BDG, we sought to understand potential sources of plasma BDG and whether it is derived from intestinal or respiratory tract fungi. To that end, we examined associations between plasma BDG with biomarkers of epithelial barrier disruption and differences in microbial load (bacterial and fungal) in each body compartment studied by culture-independent techniques.

For the respiratory tract, plasma BDG levels were significantly associated with plasma levels of RAGE (p<0.005), a biomarker of alveolar epithelial injury. This association remained significant after adjustment for clinical variables potentially related to fungal colonization in the airways (systemic antibiotics and steroid administration on day of sampling). We directly quantified bacterial and fungal load in lower respiratory tract communities (endotracheal aspirate samples) by quantitative polymerase chain reaction (qPCR) of the bacterial 16S rRNA gene and by absolute number of reads (sequences) of the Internal Transcribed Spacer (ITS) region sequencing of the fungal genome, respectively. BDG was not significantly associated with bacterial (16S qPCR) or fungal load (number of ITS reads) in lower respiratory tract samples. However, BDG remained significantly associated with RAGE (p<0.005) after adjustment for bacterial and fungal load, indicating that epithelial permeability may be a more important contributor to BDG translocation rather than the absolute amount of fungi in the airways.

To examine the potential origin of BDG from the intestinal tract, we measured plasma levels of fatty acid binding protein-2 (FABP-2), a validated biomarker of intestinal barrier integrity (31), in a random subset of 220 patients. BDG levels were significantly associated with FABP-2 (p<0.05, Figure 3), an association that remained significant after adjustment for variables potentially related to intestinal tissue ischemia and fungal overgrowth (administration of vasopressors and systemic antibiotics at time of sampling, respectively). We did not find any significant association between plasma BDG and measurements of fungal and bacterial load in stool samples in a small subset with available data (n=16). Thus, integrative analyses of biomarkers of epithelial disruption and microbial load suggested that the possibility that BDG may translocate to the systemic circulation from both the respiratory and gastrointestinal tract.

### Longitudinal evolution of BDG levels and association with outcomes

For 156 patients (34% of the cohort), follow-up plasma samples were obtained in the time window from day 3 to 5 from baseline while the patients remained in the ICU. Patients with a negative baseline test who developed a positive test on follow-up (N = 10, 6%) and those with an initial positive test who had a subsequent negative test (N = 17, 11%) had worse 30-day mortality compared to patients who remained negative on follow-up (p<0.05, Figure 5).

**Figure 4:**
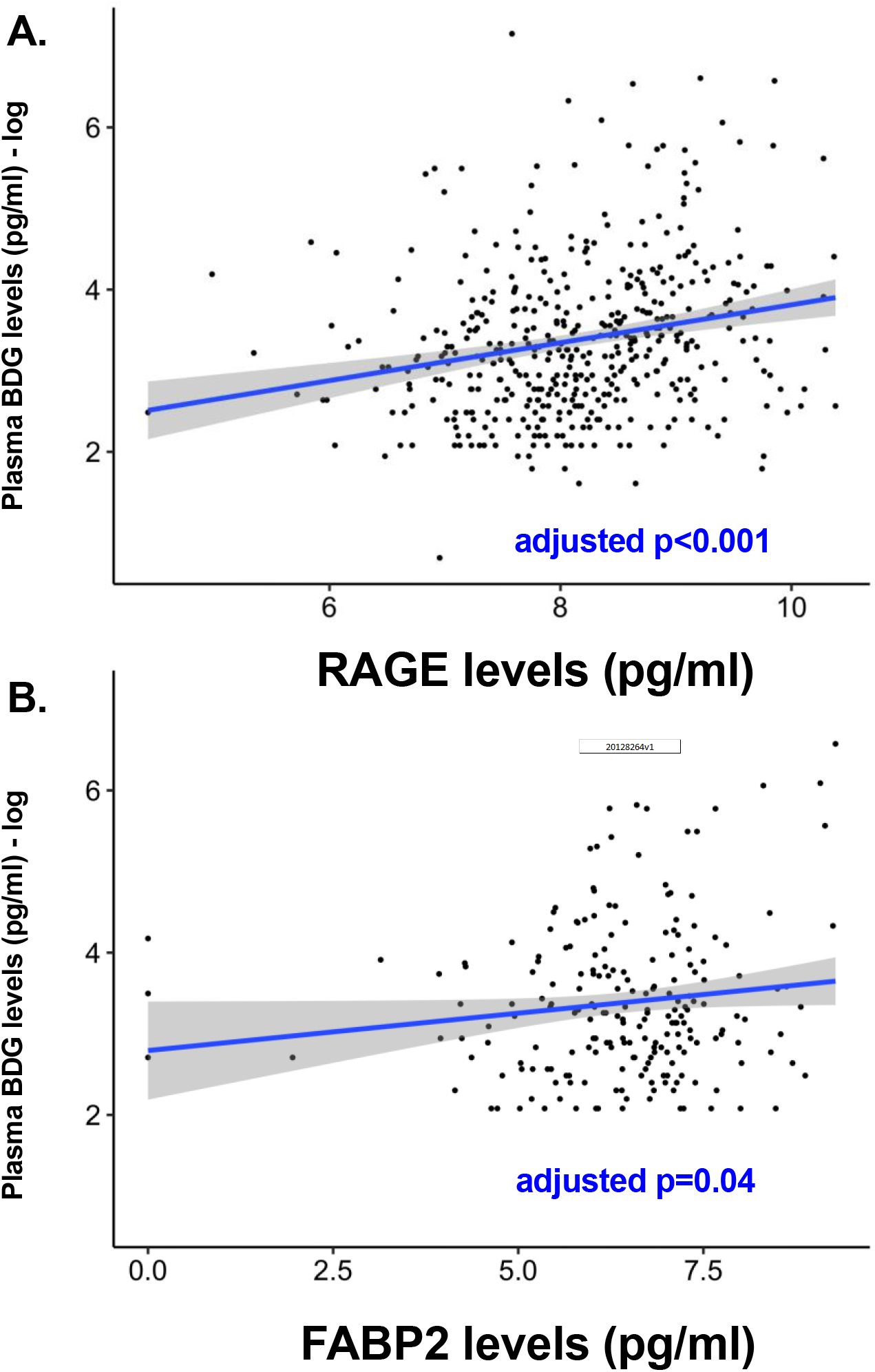
BDG levels are significantly associated with levels of markers of lung (A) and intestinal (B) epithelial barrier disruption. P-values of linear regression models are shown (A: effect of RAGE adjusted for systemic antibiotics and systemic steroids expressed as prednisone equivalent dose at the time of sampling; B: effect of FABP2 adjusted for receipt of vasopressors and systemic antibiotics).

**Figure 5:**
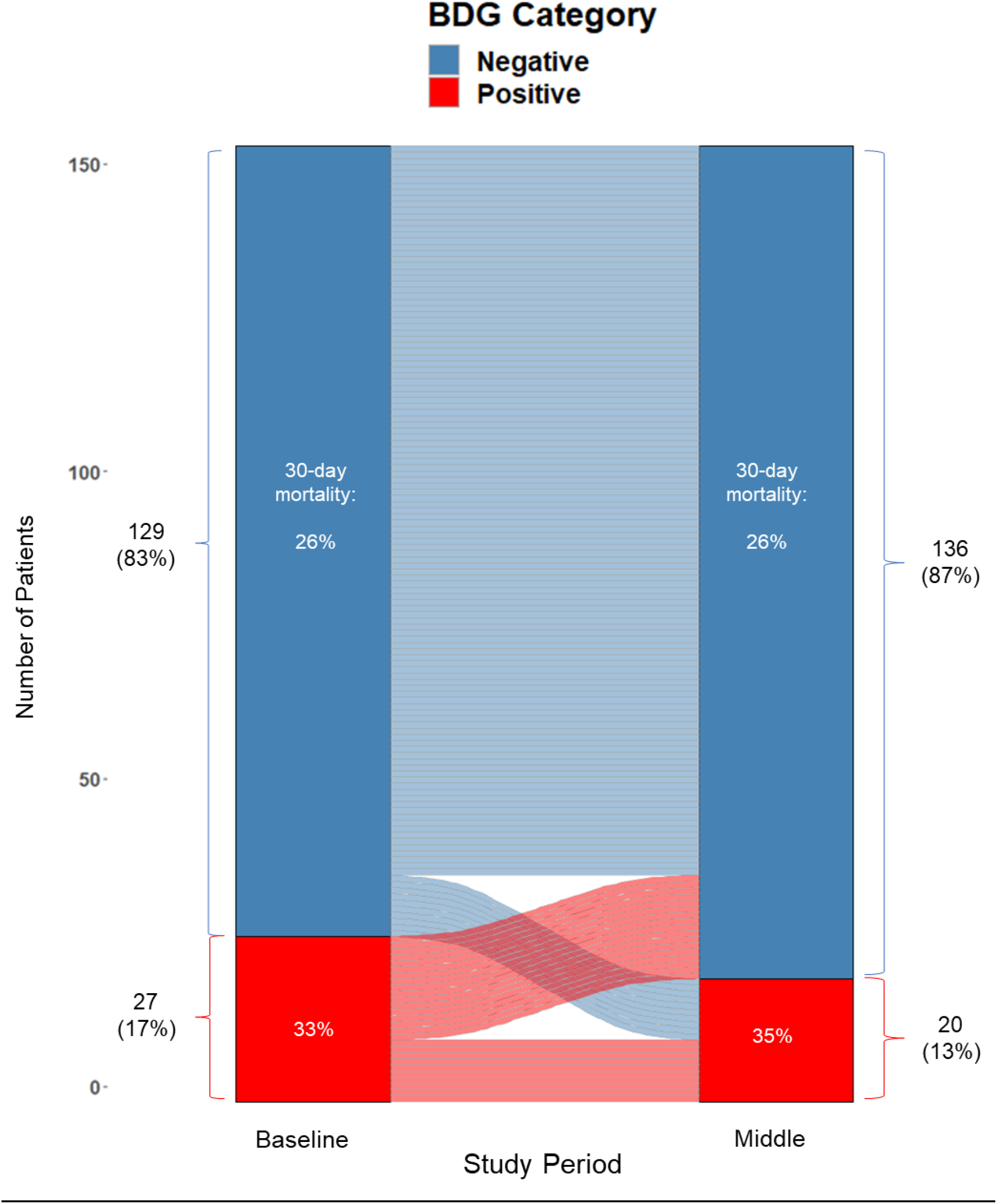
Longitudinal evolution of BDG result from baseline to 3-5 days after baseline sample and its association with 30-day mortality. Patients with a negative baseline test who developed a positive test on follow-up (N = 10, 6%) and those with an initial positive test who had a subsequent negative test (N = 17, 11%) had worse 30-day mortality compared to patients who remained negative on follow-up (60% vs 23% and 47% vs 23% respectively, p<0.05).

## Discussion

We demonstrated that BDG is detectable in the plasma in 19% of mechanically ventilated patients from a general ICU population. BDG levels correlated with host-response biomarkers of injury and inflammation and independently predicted adverse clinical outcomes. Over time, patients with incident BDG positivity had higher mortality than patients with persistently negative BDG testing. Significant associations with markers of intestinal and respiratory tract epithelial permeability suggested the possibility of fungal PAMP translocation from either vascular site, independent of fungal or bacterial loads.

BDG represents a highly immunostimulatory molecule that is likely to mediate the effects of colonizing fungi in critically ill patients. As a constituent of the cell-wall of many potentially pathogenic fungi, BDG is recognized by pattern recognition receptors such as complement receptor 3 and dectin-1 present on the surface of macrophages, dendritic cells, neutrophils and pulmonary epithelial cells (21, 32–34). Co-stimulation of dectin-1 with Toll-like receptors augments cytokine production via NF-κB-dependent (nuclear factor kappa-light-chain-enhancer of activated B cells) signaling (32, 34). Such innate immune responses, albeit necessary for the recognition and clearance of invading fungi, may also lead to exacerbated mucosal inflammation and injury upon perpetual and aberrant stimulation.

The role of BDG in the pathogenesis of mucosal injury and local inflammation, however, may not be limited necessarily to that of a direct stimulant of the innate immune system. Fungal PAMPs in the form of heat inactivated *Candida* were shown to reduce the killing activity of bone-marrow derived macrophages while enhancing cytokine production (35), while intact *Candida* has been shown to reduce the phagocytic abilities of macrophages (36), potentially by a soluble molecule (37) among other poorly understood mechanisms. Such mechanisms can lead to impaired first-line defenses of mucosal immunity with concurrent precipitation of host injury and inflammation either by fungi or other pathogenic microbes at mucosal sites. Therefore, complex interactions between colonizing fungi, non-fungal members of the microbial community and the critically-ill host need to be considered.

Dysregulation of host-responses with high levels of circulating inflammatory cytokines and immune cell activation is a key pathogenetic feature in the evolution of critical illness (38). Unsupervised classification analyses in patients with sepsis, severe pneumonia or ARDS have discovered the presence of distinct subphenotypes, characterized by a hyperinflammatory vs. a hypoinflammatory cluster of host-responses(3–5). The markedly worse outcomes and differential treatment responses associated with the hyperinflammatory subphenotype offer a new framework for predictive enrichment and the delivery of targeted efficacious therapies (e.g. anti-inflammatory drugs to be given only to patients with the hyperinflammatory subphenotype who are likely to benefit) (6). However, the proximal, molecular determinants of ARDS and sepsis subphenotypes have not been defined, and, similarly, clinical stratifications by types of insult (e.g. pneumonia or extrapulmonary infection etc.) have not been informative for distinguishing host-responses (3, 39). Thus, we posit that at least some of the observed host-response heterogeneity may be accounted by host-microbial interactions. Beyond the well-recognized importance of bacterial pathogens, translocating fungal PAMPS and their interactions with innate immune cells may also significantly contribute to the evolution and outcome of critical illness.

Epidemiologic observations support the role of fungi in critical illness, even outside the context of conventionally defined IFI. Evidence of *Candida* colonization in the airways or fungal overgrowth in the gastrointestinal tract has been associated with worse clinical outcomes, including but not limited to the development of serious bloodstream infections due to systemic invasion of fungi from different mucosal surfaces (16, 17, 19, 20). Factors that have been associated with fungal colonization in critically-ill patients include widespread antibiotic use, which allow fungi to grow and overtake environmental niches (20), as well as primary or iatrogenic immunosuppression, such as with inhaled corticosteroid therapies leading to upper respiratory tract fungal colonization (40). Our study utilized culture-independent methods (BDG measurement and next-generation sequencing approaches) to examine for translocation of fungal elements into the blood stream, their interactions with innate immunity and prognostic importance in a broad ICU population with no clinical evidence of IFI.

We observed associations of circulating BDG with innate immune responses as well as biomarkers of epithelial permeability in the respiratory and intestinal tract. BDG may be stimulating innate immunity not only through mucosal interactions, but also in the bloodstream and in tissue locations within the reticuloendothelial system (35, 41). Therefore, circulating BDG molecules may not simply represent a marker of mucosal barrier injury that has found its way into the bloodstream, but also an active mediator of inflammatory organ dysfunction and illness severity in sepsis (9, 35). Our integrative analyses of permeability biomarkers and culture-independent assays of bacterial/fungal load did not reveal a specific primary source of circulating BDG from either the lungs or the gut. However, we identified significant independent associations of BDG with corresponding markers of epithelial permeability in both compartments. With regards to the lungs, soluble RAGE, an extensively validated biomarker of ARDS radiographic and clinical severity (25, 42, 43), was strongly associated with plasma BDG levels, potentially reflecting the degree of pulmonary epithelial/endothelial disruption and ability of BDG from airway colonizing fungi to enter the systemic circulation. Similarly for the gut, BDG levels were also associated with FABP2, a biomarker of intestinal barrier disruption (31). In animal models of intestinal leakage via direct physical or chemical injury, serum BDG testing showed low sensitivity but high specificity for a permeable gut (44). Direct measurement and study of the phenomenon of microbial translocation in human translational studies is challenging, and no gold-standard method exists for *in vivo* study (45). Thus, blood levels of BDG may offer a non-invasive and clinically available surrogate measure of gut/lung permeability in critically ill patients, with important prognostic implications.

A positive BDG test (defined as >60pg/ml) conferred an adverse prognosis in our inclusive cohort of subjects with acute respiratory failure. BDG-positive patients demonstrated higher severity of illness by SOFA score, accumulated more severe organ dysfunction (incident AKI and fewer VFDs) and had significantly worse 30-day survival. These associations with adverse outcomes remained significant after multivariate model adjustments for age, severity of illness and host-response subphenotypes, suggesting that BDG testing may capture prognostic information not offered by clinically available and emerging stratification tools based on biomarkers. Similar observations of poor prognosis associated with BDG had been reported in smaller cohorts of patients with suspected VAP as well as in patients with sepsis (23, 44). Our findings in a much larger and inclusive cohort suggest that BDG may offer additional prognostic information in the course of critical illness. Further validation of the prognostic utility of BDG in independent cohorts is needed.

Our study has several limitations. It is a single-center study, and the generalizability of our findings in critically ill populations beyond our tertiary care ICU requires external validation. Our study is also limited by the available sample size. Despite being the largest study of BDG measurement in acute respiratory failure, results from analyses for subgroups and specific biomarkers (e.g. FABP2) require cautious interpretation, as the effective sample size for certain analyses was smaller. Although we aimed to control for potential confounders of BDG measurement from available variables (batch of measurement, beta-lactam antibiotics or bacteremia), there may be other sources of BDG false positivity, such as hemodialysis or blood product preparation filter processing that were not possible to account for in our analyses (46, 47). Finally, the extent of microbiologic workup for diagnosis of bacterial infection or IFI was directed by the treating physicians and, as such, microbiologic testing was not uniform. Thus, it is possible that some of the BDG positive cases may represent clinically undiagnosed or preclinical IFI. Nonetheless, inclusion of patients with IFI in sensitivity analyses did not impact the observed associations.

In summary, we demonstrate that circulating BDG is an independent predictor of a hyperinflammatory host-response profile and adverse clinical outcomes in mechanically ventilated patients with acute respiratory failure. These associations highlight a novel, underappreciated mechanism of biological heterogeneity in critical illness that involves interactions between translocating fungal PAMPs and innate immunity, which is independent of fungal load at mucosal sites and points towards complex interactions between the host, fungi and other microbes. Our findings call for further mechanistic interrogation in animal studies and independent prospective clinical study to validate the biologic relevance and prognostic information of circulating BDG.

## Methods

### Clinical cohort and sample collection

From October 2011 – June 2019, we prospectively enrolled a convenience sample of consecutive, adult patients with acute respiratory failure, who were intubated and mechanically-ventilated in the Medical or Cardiac Intensive Care Units (ICU) at the University of Pittsburgh Medical Center. Exclusion criteria included inability to obtain informed consent, presence of tracheostomy, or mechanical ventilation for more than 72 hours prior to enrollment. The study was approved by the University of Pittsburgh Institutional Review Board (protocol PRO10110387), and written informed consent was provided by all participants or their surrogates.

From enrolled subjects, we collected blood samples for centrifugation and separation of plasma and serum. We also collected noninvasive biospecimens for study of lower respiratory tract microbiota with endotracheal aspirates (ETA) and we analyzed stool samples for study of the intestinal microbiota, as previously described (48, 49).

### Laboratory analyses

We measured plasma BDG levels using the commercially available Fungitell® Limulus Amebocyte Lysate (LAL) assay (Associates of Cape Cod, Inc, East Falmouth, MA, USA) at the manufacturer’s facility (50). As a biomarker of intestinal permeability, we measured levels FABP2 with the Quantikine® Human FABP2/I-FABP Immunoassay (R&D Systems, Minneapolis, USA). For profiling host-responses, we constructed a custom Luminex multi-analyte panel (R&D Systems, Minneapolis) targeting biomarkers associated with ARDS outcomes (RAGE, IL-6, IL-8, IL-10, TNFR1, ST-2, fractalkine, angiopoietin-2, procalcitonin and pentraxin-3), as previously described (3). We extracted microbial DNA directly from samples and quantified bacterial load profiling by amplifying via quantitative polymerase chain reaction the V3-V4 hypervariable region of the bacterial 16S rRNA gene as previously described (48). For fungal community profiling, we sequenced amplified regions 1 and 2 of the ITS rRNA gene on the Illumina MiSeq platform (Supplement).

### Clinical classifications

A consensus committee reviewed clinical and radiographic data and performed retrospective classifications of the etiology and severity of acute respiratory failure. Classifications were performed without knowledge of biomarker data. We retrospectively classified subjects as having ARDS per established criteria (49), at risk for ARDS due to the presence of direct (pneumonia or aspiration) or indirect (e.g. extra-pulmonary sepsis or acute pancreatitis) lung injury risk factors (51) but lacking ARDS diagnostic criteria, acute respiratory failure without risk factors for ARDS, or acute on chronic respiratory failure. We recorded clinical microbiologic results of respiratory and blood culture specimens obtained within 48hrs of research sample acquisition. We excluded patients with diagnosis of IFI based on microbiologic evidence and consistent clinical picture (24). We recorded antibiotic prescriptions, including beta-lactams associated with false-positive BDG tests, as well as topical antifungals for skin candidiasis or oral thrush. We followed patients prospectively for incidence of AKI, VFD at 30 days, and 30-day mortality.

### Statistics

From baseline plasma BDG levels, we classified patients into those with a negative (≤60 pg/ml) or positive (>60 pg/ml) result. In sensitivity analyses, we also classified BDG levels using an 80pg/ml threshold(52). Biomarker values were log-transformed for use in regression models. From available biomarker and clinical variables, we classified patients into hyper-inflammatory vs. hypo-inflammatory subphenotype based on an internally derived and validated parsimonious logistic regression model, following application of latent class analyses (53). Baseline characteristics and outcomes between groups were compared using Wilcoxon rank-sum for non-normally distributed continuous variables and Student’s t-test for normally distributed continuous variables. Fisher’s exact was used for categorical variables. We used linear regression models to examine for associations between BDG levels and biomarkers of host responses. Multiple testing adjustments were implemented with the Benjamini-Hochberg method. We examined for differences in BDG levels between hyper-inflammatory and hypo-inflammatory subphenotypes using Wilcoxon rank sum tests. We examined for associations between BDG levels and clinical outcomes with logistic regression models for AKI and 30-day mortality, zero-inflated negative binomial models for VFD, and Cox-proportional hazard models for 30-day survival, adjusted for age, SOFA score, subphenotype classification, beta-lactam antibiotics and batch of BDG measurement. We examined for association between BDG levels and RAGE (marker of alveolar epithelial damage) using a linear regression adjusted for variables potentially associated with fungal overgrowth in the airways (systemic antibiotics and systemic steroids expressed as prednisone equivalent dose at the time of sampling). For the association between BDG and FABP2 (marker of intestinal permeability), we built a linear regression model adjusted for variables that may impact the integrity of the intestinal epithelium (receipt of vasopressors) and fungal overgrowth in the gut (systemic antibiotics). For bacterial and fungal microbiota analyses, we performed comparisons of microbial load (16S qPCR and N of fungal ITS reads) by BDG test results. Statistical significance was defined as p<0.05. All statistical analyses were performed with the R statistical software, version 3.6.0 (54).

### Study approval

The University of Pittsburgh Institutional Review Board approved the study (STUDY19050099) and written informed consent was provided by all participants or their surrogates in accordance with the Declaration of Helsinki.

## Data Availability

The data that support the findings will be available on Github at https://github.com/MicrobiomeALIR following acceptance of this manuscript for publication.

## Author contributions

**Georgios D. Kitsios:** Conceptualization, Methodology, Investigation, Software, Formal analysis, Writing - Original Draft, Writing - Review & Editing, Visualization, Supervision

**Daniel Kotok:** Conceptualization, Methodology, Investigation, Software, Formal analysis, Writing - Original Draft, Writing - Review & Editing, Visualization, Supervision

**Haopu Yang:** Methodology, Investigation, Writing - Review & Editing, Visualization

**Malcolm Finkelman:** Methodology, Investigation, Writing - Review & Editing

**Yonglong Zhang:** Investigation, Writing - Review & Editing

**Noel Britton:** Investigation, Writing - Review & Editing

**Rui Guo:** Investigation, Writing - Review & Editing

**John W. Evankovich:** Investigation, Writing - Review & Editing

**William Bain:** Investigation, Writing - Review & Editing

**Faraaz Shah:** Investigation, Writing - Review & Editing

**Yingze Zhang:** Investigation, Writing - Review & Editing

**Panayiotis V. Benos:** Methodology, Investigation, Software, Writing - Review & Editing, Visualization

**Bryan J. McVerry:** Conceptualization, Methodology, Investigation, Writing - Original Draft, Writing - Review & Editing, Supervision

**Alison Morris:** Conceptualization, Methodology, Investigation, Writing - Original Draft, Writing - Review & Editing, Supervision

## Acknowledgements

We would like to thank all members of the research team of the Acute Lung Injury Registry (ALIR) and Biospecimen Repository at the University of Pittsburgh, the medical and nursing staff in the Medical Intensive Care Unit at the University of Pittsburgh Medical Center, and all patients and their families for participating in this research project. Off-label use disclaimer: “Fungitell^®^ is cleared for use as an adjunct test in the diagnosis of invasive fungal disease. The data presented herein are for research purposes only.”

## List of abbreviations

ARDS: Acute Respiratory Distress Syndrome
ALIR: Acute Lung Injury and Biospecimen Repository
AKI: Acute kidney injury
CHF: Congestive heart failure
ICU: Intensive care unit
IL: Interleukin
ITS: Internal transcribed spacer
IFI: Invasive fungal infection
P:F ratio: Ratio of partial pressure of arterial oxygen [P] and fraction of inspired oxygen [F]
RAGE: Receptor of advanced glycation end-products
SOFA: Sequential Organ Failure Assessment
ST-2: Suppression of tumorigenicity-2
TNFR1: Tumor necrosis factor receptor 1
UPMC: University of Pittsburgh Medical Center
VFD: Ventilator-free days

## Bibliography

1. Hotchkiss RS, Sherwood ER. Getting sepsis therapy right: Is decreasing inflammation or increasing the host immune response the better approach? Science (80-) 2015;347:1201–1202.

2. Englert JA, Bobba C, Baron RM. Integrating molecular pathogenesis and clinical translation in sepsis-induced acute respiratory distress syndrome. JCI Insight 2019;

3. Kitsios GD et al. Host-Response Subphenotypes Offer Prognostic Enrichment in Patients With or at Risk for Acute Respiratory Distress Syndrome. Crit Care Med 2019;1.doi:10.1097/CCM.0000000000004018

4. Sinha P et al. Development and validation of parsimonious algorithms to classify acute respiratory distress syndrome phenotypes: a secondary analysis of randomised controlled trials. Lancet Respir Med 2020;8:247–257.

5. Seymour CW et al. Derivation, validation, and potential treatment implications of novel clinical phenotypes for sepsis. J Am Med Assoc 2019;321:2003–2017.

6. Matthay MA, McAuley DF, Ware LB. Clinical trials in acute respiratory distress syndrome: challenges and opportunities. Lancet Respir Med 2017;5:524–534.

7. Vance RE, Isberg RR, Portnoy DA. Patterns of pathogenesis: discrimination of pathogenic and nonpathogenic microbes by the innate immune system. Cell Host Microbe 2009;6:10–21.

8. Newton K, Dixit VM. Signaling in innate immunity and inflammation. Cold Spring Harb Perspect Biol 2012;4:.

9. Amornphimoltham P, Yuen PST, Star RA, Leelahavanichkul A.Gut Leakage of Fungal-Derived Inflammatory Mediators: Part of a Gut-Liver-Kidney Axis in Bacterial Sepsis. Dig Dis Sci 2019;64:2416–2428.

10. O’Dwyer DN et al. Lung microbiota contribute to pulmonary inflammation and disease progression in pulmonary fibrosis. Am J Respir Crit Care Med 2019;199:1127–1138.

11. Kitsios GD et al. Dysbiosis in the intensive care unit: Microbiome science coming to the bedside. J Crit Care 2017;38:84–91.

12. Kitsios G et al. The Human Microbiome in Critical-Illness: Longitudinal Evolution of Oral, Lung and Gut Communities in Mechanically-Ventilated Patients and Association with Clinical Outcomes. At <www.atsjournals.org>.

13. McDonald D et al. Extreme Dysbiosis of the Microbiome in Critical Illness. mSphere 2016;1:e00199–16.

14. Watkins RR et al. Admission to the intensive care unit is associated with changes in the oral mycobiome. J Intensive Care Med 2017;32:278–282.

15. Tipton L, Ghedin E, Morris A. The lung mycobiome in the next-generation sequencing era. Virulence 2017;8:334–341.

16. Pendleton KM et al. Respiratory Tract Colonization by Candida species Portends Worse Outcomes in Immunocompromised Patients. Clin Pulm Med 2018;25:197–201.

17. Delisle M-S et al. The clinical significance of Candida colonization of respiratory tract secretions in critically ill patients. J Crit Care 2008;23:11–17.

18. Delisle M-S et al. Impact of Candida species on clinical outcomes in patients with suspected ventilator-associated pneumonia. Can Respir J 2011;18:131–136.

19. Samonis G et al. Levofloxacin and Moxifloxacin Increase Human Gut Colonization by Candida Species. Antimicrob Agents Chemother 2005;49:5189–5189.

20. Vardakas KZ et al. Candidaemia: incidence, risk factors, characteristics and outcomes in immunocompetent critically ill patients. Clin Microbiol Infect 2009;15:289–292.

21. Calabrese DR et al. Dectin-1 genetic deficiency predicts chronic lung allograft dysfunction and death. J Clin Investig Insight 2019;4:.

22. Mittal R, Coopersmith CM. Redefining the gut as the motor of critical illness. Trends Mol Med 2014;20:214–223.

23. Heyland D, Jiang X, Day AG, Laverdiere M. Serum β-d-glucan of critically ill patients with suspected ventilator-associated pneumonia: Preliminary observations. J Crit Care 2011;26:536.e1-536.e9.

24. Hage CA et al. Microbiological laboratory testing in the diagnosis of fungal infections in pulmonary and critical care practice. an official american thoracic society clinical practice guideline. Am J Respir Crit Care Med 2019;200:535–550.

25. Kotok D et al. The evolution of radiographic edema in ARDS and its association with clinical outcomes: A prospective cohort study in adult patients. J Crit Care 2020;doi:10.1016/j.jcrc.2020.01.012

26. Ito S et al. False-positive elevation of 1,3-beta-D-glucan caused by continuous administration of penicillin G. J Infect Chemother 2018;24:812–814.

27. Mennink-Kersten MASH, Warris A, Verweij PE. 1,3-β-D-Glucan in Patients Receiving Intravenous Amoxicillin–Clavulanic Acid. N Engl J Med 2006;354:2834–2835.

28. Sedgewick AJ, Shi I, Donovan RM, Benos P V. Learning mixed graphical models with separate sparsity parameters and stability-based model selection. BMC Bioinformatics 2016;17:S175.

29. Sedgewick AJ et al. Mixed graphical models for integrative causal analysis with application to chronic lung disease diagnosis and prognosis. Bioinformatics 2019;35:1204–1212.

30. Liu Q et al. IL-33–mediated IL-13 secretion by ST2+ Tregs controls inflammation after lung injury. JCI Insight 2019;at <https://insight.jci.org/articles/view/123919>.

31. Shieh A, Epeldegui M, Karlamangla AS, Greendale GA. Gut permeability, inflammation, and bone density across the menopause transition. J Clin Investig Insight 2020;5:.

32. Schorey JS, Lawrence C. The pattern recognition receptor Dectin-1: from fungi to mycobacteria. Curr Drug Targets 2008;9:123–129.

33. Camilli G, Tabouret G, Quintin J. The Complexity of Fungal β-Glucan in Health and Disease: Effects on the Mononuclear Phagocyte System. Front Immunol 2018;9:673.

34. Brown GD et al. Dectin-1 is a major beta-glucan receptor on macrophages. J Exp Med 2002;196:407–412.

35. Panpetch W et al. Gastrointestinal Colonization of Candida Albicans Increases Serum (1→3)-β-D-Glucan, without Candidemia, and Worsens Cecal Ligation and Puncture Sepsis in Murine Model. Shock 2018;49:62–70.

36. Seider K et al. The Facultative Intracellular Pathogen Candida glabrata Subverts Macrophage Cytokine Production and Phagolysosome Maturation. J Immunol 2011;187:3072–3086.

37. Collette JR, Zhou H, Lorenz MC. Candida albicans Suppresses Nitric Oxide Generation from Macrophages via a Secreted Molecule. In: Chauhan N, editor. PLoS One 2014;9:e96203.

38. Meyer NJ, Calfee CS. Novel translational approaches to the search for precision therapies for acute respiratory distress syndrome. Lancet Respir Med 2017;5:512–523.

39. Calfee CS et al. Subphenotypes in acute respiratory distress syndrome: latent class analysis of data from two randomised controlled trials. Lancet Respir Med 2014;2:611–620.

40. Rachelefsky GS, Liao Y, Faruqi R. Impact of inhaled corticosteroid-induced oropharyngeal adverse events: results from a meta-analysis. Ann Allergy, Asthma Immunol 2007;98:225–238.

41. Yoshida M et al. Detection of plasma (1 --> 3)-beta-D-glucan in patients with Fusarium, Trichosporon, Saccharomyces and Acremonium fungaemias. J Med Vet Mycol 35:371–4.

42. Kitsios GD et al. Host-Response Subphenotypes Offer Prognostic Enrichment in Patients With or at Risk for Acute Respiratory Distress Syndrome. Crit Care Med 2019;47:1724–1734.

43. Mrozek S et al. Elevated Plasma Levels of sRAGE Are Associated With Nonfocal CT-Based Lung Imaging in Patients With ARDS: A Prospective Multicenter Study. Chest 2016;150:998–1007.

44. Leelahavanichkul A et al. Gastrointestinal Leakage Detected by Serum (1→3)-β-D-Glucan in Mouse Models and a Pilot Study in Patients with Sepsis. SHOCK 2016;46:506–518.

45. Deitch EA. Gut-origin sepsis: evolution of a concept. Surg 2012;10:350–356.

46. Nagasawa K et al. Experimental proof of contamination of blood components by (1?3)-?-D-glucan caused by filtration with cellulose filters in the manufacturing process. J Artif Organs 2003;6:49–54.

47. Kanda H et al. Influence of various hemodialysis membranes on the plasma (1→3)-β-D-glucan level. Kidney Int 2001;60:319–323.

48. Kitsios GD et al. Respiratory Microbiome Profiling for Etiologic Diagnosis of Pneumonia in Mechanically Ventilated Patients. Front Microbiol 2018;9:1413.

49. Fair K et al. Rectal Swabs from Critically Ill Patients Provide Discordant Representations of the Gut Microbiome Compared to Stool Samples. mSphere 2019;4:e00358–19.

50. 1,3-beta-D-glucan Fungitell ®. at <https://www.fungitell.com/pdfs/Fungitell_Brochure_PR18-016web.pdf>.

51. Neto AS et al. Epidemiological characteristics, practice of ventilation, and clinical outcome in patients at risk of acute respiratory distress syndrome in intensive care units from 16 countries (PRoVENT): an international, multicentre, prospective study. Lancet Respir Med 2016;4:882–893.

52. Caporaso JG et al. Ultra-high-throughput microbial community analysis on the Illumina HiSeq and MiSeq platforms. ISME J 2012;6:1621–1624.

53. McMurdie PJ, Holmes S. phyloseq: an R package for reproducible interactive analysis and graphics of microbiome census data. PLoS One 2013;8:e61217.

54. R Core Team. R: A Language and Environment for Statistical Computing. 2018;at <https://www.r-28project.org/ >.

